# Dehumanisation and Datafication: Key stressors of Algorithmic Management: A qualitative analysis of Chinese couriers

**DOI:** 10.1101/2022.09.13.22279887

**Authors:** Hua Wei, Huiyue Shi, Yilin Kou, Mengke Yu, Shugang Li, Thomas O’Toole, Christopher J. Armitage, Tarani Chandola, Pauline Whelan, Yanchun Zhang, Yan Xu, Martie van Tongeren

**Author notes:** Correspondence: Hua Wei.

## Abstract

**Introduction:** Technology is an important social determinant of health that has so far been poorly understood. Nevertheless, technologies such as algorithms and artificial intelligence are rapidly changing how work is organised and managed globally. Courier work is one of the most affected occupations due to the common use of digital labour platforms. This study will explore how AM shapes Chinese couriers’ work experience and engenders health inequity.

**Method:** We conducted 15 in-depth interviews with couriers in May-June 2021 in China. Thematic analysis was completed using Nvivo 12.

**Results:** AM can introduce new work stressors and interact with existing factors to alleviate or intensify work stress. Couriers’ responses to AM and its effects varied as some felt motivated while others frustrated. Key themes emerged from the interviews included ‘App the “boss”’, ‘Unequal exposure to AM and the role of human support’, ‘New work stressors of AM – Dehumanisation and Datafication’, ‘Job stressors intensified by AM’ and ‘Varying individual response to AM’. App ‘the boss’ described how the couriers felt when managed by algorithms. Employed, gig and agency couriers had different exposure to AM and human support of different quality. Dehumanisation characterizes how algorithmic systems fulfil managerial functions in ways that differ from managers qualitatively or quantitatively. Datafication refers to the conversion of real-world information and user behaviour into data and being input in the system to assist automated decision-making. Couriers also reported that the use of digital methods could intensify some work stressors, e.g. time pressure and customer behaviour. Nevertheless, some couriers reported that AM provided new job resources, e.g. flexible hours, job opportunities, pay transparency, and bypassing office politics.

**Conclusion:** This paper identified key work stressors of AM and explored how AM interacts with existing work stressors to shape Chinese couriers’ work experience. Dehumanisation could reduce the quality of support and their sense of relatedness to the workplace. Datafication or extraction of data from workers and lack of transparency led to concerns about potential discrimination, workplace unfairness and power imbalance. Couriers’ responses to AM varied and future research should take such heterogeneity into account.

## 1 Introduction

Despite the increasing prevalence of managerial algorithms and artificial intelligence (AI) at work in all sectors, our understanding about how technology shapes worker experience and engenders or reinforces health inequity is limited ^1^. Algorithmic Management (AM) has been defined by Möhlmann, Zalmanson ^2^ as: “the large-scale collection and use of data on a platform to develop and improve learning algorithms that carry out coordination and control functions traditionally performed by managers”. It reveals three key elements of AM, data inputs, system logics and management outputs ^3,4^. In addition to the data process that executes managerial functions, it also highlights the key feature of AM that is the substitution of managers ^5^. This is an essential part of digitalization that revolutionizes the workplaces beyond the gig economy due to high replicability and scalability ^6–9^. However, existing research about algorithmic management (AM) has predominantly focused on its functionalities and failed to capture the underlying mechanism of AM as a potential source of work stress. In other words, the focus has been on “what does AM manage”, that is essentially the same to human managers, but not on “how does AM manage”, where the difference lies between AM and human managers. Finding answers to the “how” question necessitates qualitative, exploratory work that examines the process of AM ^10^.

Although AM is transforming almost every sector of the economy, the logistics sector is one of the most advanced and couriers are the key workforce in this sector ^11^. This is due to the expansion of digital labour platforms, such as Uber and Deliveroo ^12–14^, or their equivalents in China: Meituan, Ele.me and Didi ^15,16^. Working conditions of couriers, particularly in China, appear to be highly stressful and unsafe ^17,18^. Since the beginning of the COVID-19 pandemic, media reports of sudden death of young couriers due to overwork emerged in South Korea, China, and Singapore (See for example, <u>Delivery workers in South Korea say they’re dying of ‘overwork’</u> (sanjuandailystar.com)). Karoshi or sudden death from overwork is often caused by stroke, resulted from a repeated triggering of stress responses ^19^. Lin et al developed a couriers’ overwork scale to investigate risks of Karoshi among Chinese takeaway and parcel couriers. They found that among takeaway couriers, about 41.7% of respondents (n=1114) were at high risk of overwork, 35.9% were at very high risk and only 7.9% were considered risk-free ^20^. The situation was similarly severe among parcel couriers (n=1214), where 40.1% were at high risk, 38.6% were at very high risk and only 3.7% were risk-free^21^. Road safety is another major concern among couriers. A recent cross-sectional study of road safety situations among Chinese parcel and food couriers revealed that 76.5% of interviewed couriers (n=480) had been involved in a traffic crash at least once, despite the average length of staying in this occupation was only about 1.5 years ^18^. The same study also conducted 600 roadside observations and found that traffic violations were very common, such as speeding, riding in the motor vehicle lane, and use of a cell phone whilst riding. The authors believed that the tight delivery schedule set by the platforms was one of the main reason for these traffic violations. Other studies found that high job demands, long working hours etc among Chinese couriers are associated with poor job satisfaction and health outcomes ^22,23^. The socioeconomic status of informal, migrant workers in China’s urban area may further exacerbate health inequity among the couriers ^24,25^. Although AM was not explicitly considered in these studies, the authors highlighted the potential impact of digitalized work arrangement and insecure employment status.

The situation is alarming, suggesting Chinese couriers may be at high risk of worsened work environment driven by AM. Nevertheless, health researchers are not equipped to measure occupational risks against AM due to the absence of a working definition of the key stressors and measurement. Research that have made attempts to measure AM [e.g. Cram, Wiener ^26^, Sun ^27^, and Rani, Pesole ^28^] have either avoided defining work stressors inherent to the process of AM or ignored the underlying mechanism of health impacts. ^3,29^. It must be recognized that AM reverses the relationship between artefacts and users ^30^. In other words, our understanding about the role of technology and how it impacted people’s life were based on the assumption that the technology was a tool without learning capacity. When AM begins to substitute managers and make decisions about people, it has become ‘the boss’ ^5,31^. It creates a new path of impact and renders models built on the old assumptions obsolete ^30^. Hence, it is necessary to explore from the workers’ perspective, or those ‘being managed’, to understand whether AM has introduced new work stressors that differ from stressors under human management. This research set out to improve our understanding about the underlying mechanisms of AM in relation to health and safety outcomes among couriers in China.

## 2 Materials and methods

We took a qualitative and explorative approach to address the research questions.

### 2.1 Data collection

We conducted 15 semi-structured interviews in May and June 2021 in China using Tencent Meeting software. Each of the interviews lasted about 90 minutes. We approached potential participants through snowballing, started by approaching couriers working in the researchers’ vicinity. About half of those we contacted refused to participate mainly due to being too busy. Recruitment stopped when the seven interviewers agreed that data saturation was reached. All interviews were carried out in Chinese.

The study was reviewed and approved by ethics committees in both the UK and China (University of Manchester Research Ethics Committee Ref: 2021-8761-18609 and Beijing Normal University Ethics Review Committee, IRB No.: 20200113001). All methods were performed in accordance with the relevant guidelines and regulations. Each participant received a Participant Information Sheet and informed consent was confirmed either by verbal consent at the beginning of the session or by written consent. Compensation were provided for participants’ time. Researchers introduced themselves and the research purpose to the interviewees. Seven researchers conducted the interviews, with each session completed by at least two researchers. All researchers were trained for qualitative research. Two interviewers have PhD degrees and the other researchers were postgraduate students. Four of the interviewers were female. Interview questions included open inquiries about work stress, individual response and coping methods. The role of technology was elicited or probed during the conversation to allow the interviewees form their perspectives and makes sense of the phenomenon ^10,32^. The interview schedule is available in Supplementary file.

### 2.2 Data analysis

We followed the consolidated criteria for reporting qualitative studies (COREQ) to report our findings (Tong et al., 2007). A reporting checklist can be found in Supplementary file. Data were analysed in Chinese, with quotes translated into English by the bilingual researchers. Coding was completed using Nvivo12 software, with codes generated inductively (Braun & Clarke, 2006; Castleberry & Nolen, 2018). Researchers first pilot coded three transcripts to generate codes. Relevant codes were grouped to form themes and the initial codebook. The initial codebook was discussed with the rest of the research team and revised based on the feedback. The researchers then coded all the transcripts based on the final codebook. Five researchers each coded two same transcripts independently. Coding results were merged to assess inter-coder reliability. The average Kappa coefficient of each coder against the second coder worked on the same documents ranged from 0.54 to 0.63 (0.41–0.75 is considered fair to good ^33^). The percentage of agreement between the two coders was generally very high (>90%). Disagreements were discussed until consensus reached.

## 3 Results

Of the 15 interviewees, 11 were Beijing-based, two based in a second-tier city and another two based in a smaller city. Two of the interviewees were female and 13 were male. The mean age was 33 (22-57 years old) and average length of working as a courier was 3.5 years (2 months-9 years). We interviewed six parcel couriers and nine takeaway food couriers. There were three employment types, directly employed, agency worker and gig worker. We interviewed six employed couriers (0509e, 0523e, 0524e, 0529e, 0601e, 0607e), three agency couriers (0525a, 0528s, 0529a) and six gig couriers (0510g, 0510-Yg, 0517g, 0518g, 0522g, 0604g). Employed couriers were all parcel couriers.

### 3.1 App ‘the boss’

**App ‘the boss’** describes how the interviewees felt when managed by algorithms. The app or the system functioned like a manager and dictated who got the job, and how the job should be completed, evaluated and paid. One interviewee (0517g) put it simply as:

*No matter how long we have been working [for the app], a few months or a few years, we all have to listen to the system, no matter who. Nobody can interfere with the system. The system is the boss. It decides*.

Or as another interviewee (0525a) put it:

*The system determines whether you are busy or can take it easy*.

As the couriers seemingly accept the app or the system as a dictating boss, there was a strong sense of powerlessness. Some interviewees described the system just like a manager. One interviewee (0528a), for example, believed the algorithm would balance the workload for the couriers.

*The system will allocate [orders]. Where we should go or the distance of our orders would be proportionate to the total amount of orders that we deliver every month. The system will decide, based on each individual’s workload, that where we deliver to. It won’t be too far [from the supposed ratio]. It [the system] will balance the overall workload*.

Interviewees also noted the system’s learning capacity. It was believed that the system was intelligent and learning from multiple data sources, as described by an interviewee (0517g).

*The system, you can’t imagine, the system is really smart, like it knows everything. I’d say they even know whether the restaurants are slow or quick in preparing food. The system is just smart*.

Nevertheless, a key problem associated with app the boss was that although this ‘boss’ could learn from data and make decisions based on average, predictable situations, it lacked the capability to listen, process and respond to individual cases. After an interviewee (0510g) described a range of stressors, such as restaurant delays, customer behaviours and road traffic, the researcher asked whether there were any ways for the system to accommodate unusual situations or challenges beyond couriers’ control.

*They would not allow couriers to have voices (in the system). Whatever the algorithm tells you to do, you just have to do it. If you were late, penalties on you…Even the customer service would not be able to interfere [with the algorithms]. If you phone them up, and they support you, they could only put a note in so you don’t get penalties. (0510g)*

### 3.2 Unequal exposure to AM and the role of human support

Although AM techniques were commonly used but not equally distributed within the occupation. It appeared to us that employment types may be an indicator of the intensity of exposure to AM. Gig couriers appeared to be the most algorithmically managed, employed couriers appeared to be the least, with agency couriers somewhere in between. Employed couriers had employment contract with their companies and were hired or fired through the HR department. They had frontline managers and access to other parts of the organisation. They also had a depot to report to on daily basis. For employed couriers, job allocation, route planning and partially performance were managed by algorithms and their managers played an important role in mediating relations between the couriers and the organisation and sometimes customers. For example, 0607e told us how his manager mediated the relationships when he faced with aggressive customer behaviour following three failed attempts of delivery.

*When my colleague arrived at the address, the customer came out with a knife and chased him. The police was called and the site manager got involved to investigate. We had a long chat. He arranged for me to apologize and buy some small presents for the customer. I was extremely upset that I had to apologize. I was so reluctant. But the site manager said if the customer made a complaint, I would be fined for ¥2000 by the company and possibly fired. He suggested the presents would appease the customer and wouldn’t cost much*.

In this case, if the customer issued a formal complaint, automatically the courier would face a hefty fine. If the complaint was too serious, it would cost him the job. The manager provided his support based on his understanding what was the best solution for the courier. We also sensed that the courier did not like the solution as he felt it was unfair. Hence, the role of manager in this case was two-fold. On the one hand, he moderated the impact of AM by advising the courier how to avoid the fine as the system’s logic automatically equates customer complaint to fines on couriers. On the other hand, he reinforced the system logic by persuading the courier to accept a solution that was considered unfair by the courier.

Both gig couriers and agency couriers we interviewed worked for food and grocery delivery platforms. Being platform-based meant gig couriers and agency couriers were both highly algorithmically managed with limited human support. Nevertheless, there were some differences in terms of their exposure to AM. Gig couriers signed up freely on the app and were not obliged to sign in at any time. They were added to a chat group that ran by a manager who only released group notices but never replied to their enquiries nor disclosed a contact number. The only non-digitalized support gig couriers getting was through the ‘customer service hotline’ provided by the platform, which was responding to customers (who placed the orders), merchants (restaurants and shops) and couriers as they were all users of the platform.

In contrast, agency couriers had zero-hour type contracts with an agency company that was different from the name of the platform. Agency couriers had a site manager and needed to report to a depot on daily basis. They were expected to work for certain number of hours per day and number of days per week. During busy hours, they were obliged to sign in and take orders. They also needed approval from the manager if they wanted to take more leaves. Although rules were tight, an agency courier interviewee reported how her manager were considerate of health conditions.

*Yes it is required to work during weekends, all riders. But like today I took the afternoon off. It’s not too difficult for me [to ask for leave]. Because it was raining and I took a vaccination shot this morning and I can’t be rained up or catch a cold. Situations like this, if you explain to him [the manager], he will let you off. (0528a)*

In this case, the manager provided some negotiating space between the system and the agency courier. In comparison, gig couriers who did not have access to managers also did not have commitment to log in or work for set hours. In this aspect, they were genuinely their own boss.

### 3.3 New work stressors of AM – Dehumanisation and Datafication

We identified two themes as they were repeatedly brought up by the interviewees when we probed into the role of technology: Dehumanisation and Datafication

**Dehumanisation** characterizes how algorithmic systems fulfil managerial functions in ways that differ from managers qualitatively or quantitatively. Gig courier interviewees described their manager as the ***“virtual manager”*** who they had never met in person, who merely disseminated notices or information in a group chat but rarely responded to anyone in the group chat and called them using virtual numbers, meaning they would never be able to trace the manager’s number or call back (0510g, 0517g). It led to difficulties in seeking support when they had accidents, for example. Gig couriers reported they were not clear about what was covered by the insurance, or that they were not familiar with the claim procedures (0518g, 0604g).

Since the “virtual manager” was inaccessible, gig couriers turned to the platforms’ customer service hotline to gain support when faced with customer abuse, restaurants delays or other challenges at work. However, as delivery schedules were so tight, contacting customer service over the phone was considered time consuming and might cause further delay (0510g, 0517g, 0522g). As described above, customer service hotline serve all types of platform users, some couriers were concerned the hotline had too much power to decide which party was at fault for any disputes with customers and might not judge fairly. One interviewee (0517g) reported a case about customer behaviour. When the researcher asked whether he sought support from the hotline, he said:

*It won’t work. Once the customer made a complaint, or wrote a bad review, nothing works. It’s fines and money deducted (by the platform)*.

When the researcher asked whether he could appeal against customer complaints or penalties, he answered:

*Of course you can. But whether they would approve is another issue. The chance is normally small*.

The absence of manager support meant gig couriers often had to navigate the system without guidance. This led to frustration and emotional responses. One of the interviewees (0604g) told us he was screaming at the sky one night because he was so frustrated:

*… It was near the end of my shift, normally 10pm… Although they say 10pm, we could never finish at 10pm. Orders often came in near 10, like 9:50 and it could be a few kilometers away. We (fellow couriers) said we’d go for dinner together that night. So I tried to cancel, tried to transfer it to other couriers, but no way… I wanted to curse someone, but didn’t know who*.

**Datafication** refers to the conversion of real-world information and user behaviour into data and being input in the system to assist automated decision-making. Gamification, for example, is a typical example of datafication and Dehumanisation. It uses game elements to increase user interaction on a product or a software application by making it more interesting and challenging (Prabowo et al., 2019). It is commonly used to engage and motivate platform-based workers. Couriers may see gamification as a challenge or hindrance stressor. This will be elaborated with examples in later section about individual response.

Although interviewees were not data experts, they were able to understand what data means when being managed by algorithmic systems.

*That is to say they have worked for long enough to see the result, hence their data looks good. My data does not look good because I’ve only done it for a few months. Even though you’re like running all day but you can’t beat them [the more experienced ones]. They could deliver 40, 50 orders per day. You work for a whole day but could only deliver 20, 30 orders. Your data will never look as good as theirs. (0517g)*

They were also able to make sense of how the system use the data and figure out the rules embedded in the system.

*No one knows how exactly the system works. It is really not clear what the situations are. But the system appeared to be working this way: if your data is good, it gives you good orders. You get orders quickly, the orders are close to each other and easier to complete… For example, if they are Gold [grade], not like us, we’re Bronze or Silver, there can be quite a bit of difference. Imagine the system allocated six orders (to one courier), they might get places like hotels, or office buildings where you just drop the orders at the reception and the customers will come down to collect them. Whilst for someone like me [low grades, e.g. Bronze], they may send me to climb six floors, or take the lift to the 22nd floor in a residential building. You certainly cannot be as quick as them, because you have to climb up stairs, take lift, wait for lift etc. During peak hours, these take lots of time. (0517g)*

The interviewee certainly sensed the discrimination against their performance and the lack of transparency as he declared ‘no one knows how exactly the system works’. Nevertheless, they form their own strategies to deal with incidents that would affect their data. For example, one interviewee mentioned one day he was very upset because of the behaviour of a customer, he then rejected all the orders allocated to him for the rest of the day. High rejection rate would negatively affect his record or ‘his data’. He then carried on to tell us:

*You’ll just have to carry on. I just carried on delivering orders the next day. The worst would be the orders will take longer, the distance [between orders] become longer, and more difficult to complete. You’ll just have to build your data again. You re-build it with a positive attitude, if you want to continue doing it. If you want to do it do it well. (0525a)*

Performance data were undoubtedly being collected, analysed and ranked, so that the system could give them grades (gold, silver, bronze), as described by the interviewees above. The interviewees tried to make sense about the process of datafication and learn to deal with discriminating rules such as ‘good data’ is associated with ‘good orders’.

The platforms seemed be able to collect personal information from platform-based couriers without much restriction. For example, the platforms were under pressure to ensure couriers safety. Hence, couriers often received requests initiated by the app, normally within minutes or sometimes seconds of completing an order, to check whether they were in full uniform and wearing helmet. When they responded, the app would use facial recognition technology to verify their identity and took photo shots of their images (0510Yg, 0518g, 0525a). The situation is the same for both agency and gig couriers. On the contrary, the employed couriers did not have to face this. Their companies took a different approach and sent inspectors on the road to check compliance and safety behaviour (0509e, 0524e).

### 3.4 Job stressors intensified by AM

The overall work environment was stressful as one of the interviewees described there were “all sorts of triggers for mental breakdowns” (0604g). We found that AM played a role to intensify other stressors.

Time pressure was extremely high among the platform-based couriers.

*You can’t afford to be stuck. You run all the time. If one order gets stuck, all the other orders down the line will be delayed. You’d be nervous. (0525a)*

“Late delivery” is a situation when order was not delivered within the time scale set by the AM or “the system”. It was a major source of stress and mentioned 79 times by 10 interviewees (one employed and 9 platform-based), of which 78 were from platform-based couriers whose orders were allocated by the apps, with driving routes planned and delivery time set. It was believed that the algorithms were designed to push couriers to work faster (0517g, 0525a).

Customer incivility represents a major source of stress. Location tracking and customer rating landed some power to customer (who place and receive the orders) to pressurize couriers further. An interviewee (0517g) shared this experience:

*Once I was delivering an order, the next customer saw me [location on the app]. I was like a few hundred meters away from him. He saw that I was not moving for like 5 or 6 minutes. But I was just delivering the order before his. He started phoning and rushing me, saying ‘I saw your location, why were you not moving?’ I said sorry I’d be quick. After 30 seconds, he phoned again, saying ‘you need to hurry, you are not moving’. I explained and told him I’d be with him right away. He saw me [location] not moving, then phoned the third time and said he would make a complaint. The moment he hang up, I completed the other order. One minute, I was 300 meters away from him, it took me one minute to get to him. I drove so quickly, was not afraid of accidents at all. I wanted to rush and worried I might get a complaint. One complaint then it’s all wasted [as fine due to customer complaint is the worth of delivery fees from several orders]. When I handed the order to him, I sincerely apologized to him and said sorry. He took the order reluctantly and said ‘sorry meant nothing’. He then made a complaint. (0517g)*

### 3.5 Varying individual response to AM

Interviewees reported varying responses and perceptions about the same feature of AM. Using gamification as an example, some of the interviewees said they enjoyed it. One interviewee talked proudly about the day when she delivered a record number of orders and regretted that she could not achieve a higher number.

*The highest number I achieved was 92 orders [in a day]…I worked normal amount of hours [that day] but it was a festival so lots of orders…there were people who could complete 100 more orders most of the days…12 o’clock [midnight] came too early [on that day]. I was 8 orders short of 100…the system did not give me the chance [to reach 100]. It’s a shame. After summer, there are fewer orders. I only get 50 to 60 orders per day now. (0529a)*

Another interviewee (0528a) stated that her desire to win kept her in this job, and that she preferred this job to her previous office-based one, because both her physical health and sleep quality improved since she switched. On the other hand, there was an interviewee who felt they could never be as good as those more experienced couriers (see quotes from the Datafication 0517g).

Because AM made performance measures highly transparent, voluntary competition among couriers were stimulated as some would show off on social media or in group chats, inducing pressure on the other couriers.

*There are all sorts of screenshots in our [WeChat] group, sometimes it’s unit price [delivery fee goes up when they achieved certain goals], sometime it’s how much they made that day, all sorts screenshots, anyway, all sorts of show-off, all sorts of psychological attack.(0604g)*

AM and digital labour platforms undoubtedly provided more job **opportunities and flexibility.** Gig couriers found the process straightforward. They just needed to download the app, register and verify identity, and there was no contract binding except for insurance where the platform paid ¥3 daily for their safety (0517g, 0518g, 0604g). They agreed that there was autonomy and freedom but outside of the system, as described by the interviewees:

*To a certain extent, freedom means that you can decide whether to do it or not, whether to work or not to work today, but once you started working, the system dictates everything. (0517g)*

*Yes, we can reject orders but not too many. If rejected too many [orders], they [the system] will restrict orders your can get. (0510Yg)*

Most of the interviewees considered it good pay and many said that their income was higher, compared to their previous jobs. In addition, they appreciated that hard work was rewarded for example, attendance bonus, workload and service quality related bonus, and weather subsidy (additional fees they receive when delivering in bad weathers), without the need to deal with office politics (0528a). We found that younger interviewees (0604g, 0528a, 0529a; aged in their twenties) tended to have a more positive response to AM and were often motivated to be competitive in algorithm-driven incentive programs, e.g. gamification and ranking, or voluntarily engaged social networks, e.g. chat groups.

## 4 Discussion

AM is a paradigm-shifting force as it challenges our long-held beliefs about what is human ability and what is machine capability ^30^. The human-like characteristics of algorithmic systems thus created a new artefact-user relationship that necessitates new definitions and models ^4^. Coming from the worker users’ perspective, or those ‘being managed’, we identified two key stressors introduced by AM: Dehumanisation and Datafication. Dehumanisation is fundamental to AM but the withdrawal of organisational support, e.g. an inaccessible virtual manager or a generic hotline with whom the couriers had no confidence whether they would be supported, might reduce the perceived quality of support and the sense of relatedness to the digitalized workplace. Datafication or extraction of data from workers, is the foundational process of AM as the system cannot deliver management outputs without the data inputs. Features such as gamification, ranking and job allocation are based on behavioural data collected from the workers. Digital labour platforms also had the power to collect biometric data from the couriers, e.g facial imagines. The interviewees did not raise the issue of data privacy or surveillance as they accepted the fact that AM is the boss and the boss dictates. There was a strong sense of power imbalance and reluctant acceptance of unfairness and discrimination. It must be recognized that Datafication can function alone to deliver such effects without Dehumanisation. Because the lack of transparency about how workers’ data are being collected, analysed and used against them plaques the workplaces, even when decision-making remains in the hands of human managers, Datafication lends great power to them to judge those ‘being managed’ ^3,5,13,34^.

Couriers’ exposure to AM was not equally distributed cross employment types and work organisation mode. This is consistent with finding from qualitative research that compared experience of Uber drivers working in London, New York and multiple other cities ^35^. The identification of key stressors of AM can assist future study to develop a typology in relation to the extent of Dehumanisation and Datafication and the perceived effects on, for example, quality of support, sense of relatedness, data transparency, discrimination, workplace unfairness and power imbalance. In terms of Dehumanisation and quality of support, organisational support provided through human, e.g. managers and customer service hotlines, should be taken into account, as they can reinforce system rules or provide some negotiating space for worker users. These findings are consistent with literature from Human-factor in Computing Systems ^15,30,36,37^ but were not articulated to explain the mechanisms of AM for health researchers.

Another key finding of this study is the interaction between AM and other psychosocial factors, e.g. time pressure and customer behaviour. Although several studies on Chinese couriers’ occupational stress, mental health and safety have highlighted the highly stressful working conditions due to algorithmic control, they did not evaluate the role of AM in intensifying the effects on health outcomes ^16,17,23,27,38^. Future studies should take into account the interactive effects between AM and other work stress factors with consideration about what it means for workers’ health and safety outcomes.

Couriers in China are at the forefront of experiencing and coping with these new job stressors and hence provide an important starting point for conceptualizing the AM and understanding the underlying mechanisms. Similar to other gig workers in the world, Chinese couriers have also tried to figure out how the system works, what it means to their work life and have developed coping strategies (McDonald et al., 2021; Möhlmann & Za^39^lmanson, 2017). However, studies focusing solely on platform-based workers, e.g. Uber or Deliveroo, provided limited insights into how workers negotiate with automated management through the limited human support provided by the system. As we talked to couriers with varying exposure to AM, the importance of human-factor was identified.

Individuals responded to AM in different ways. For example, some participants in our study enjoyed gamification and found it a fulfilling experience, whilst some other participants expressed concerns about discrimination. Some preferred the flexibility and pay transparency provided by the system while some found navigating the system without guidance frustrating. These findings are consistent with sociological research of AM ^40–42^. However, previous studies tend to focus on either positive or negative impacts of AM without taking into account that variations due to individual circumstances. Future research should further identify individual factors that predict work experience and outcomes.

### 4.1 Limitations and future research

This is an exploratory study based on a relatively small number of interviews. The insights may be nuanced and insightful about Chinse couriers, although it is unclear to what extent the findings are generalizable to other countries. However, the results appear to be broadly in line with existing sociology and psychology literature of this topic. Future research can continue to develop the concepts of Dehumanisation and Datafication and utilize it for larger studies on AM.

## 5 Declarations

### Ethics approval and consent to participate

The research has been approved by the University of Manchester Research Ethics Committee Ref: 2021-8761-18609 and Beijing Normal University Ethics Review Committee, IRB No.: 20200113001.

### Consent for publication

Consent for publication has been obtained from the participants.

### Availability of data and materials

The datasets used and/or analysed during the current study are available from the corresponding author on reasonable request.

### Authors’ contributions

MvT led the project. MvT, HW, TC, CA, PW, YX and YCZ conceived the ideas and obtained the funding. HW, HYS, YLK, MKY and SGL collected the data and analysed the results. HW drafted the manuscript. All authors reviewed and made suggestions to different versions of the manuscript. All authors read and approved the final manuscript.

## Data Availability

All data produced in the present study are available upon reasonable request to the first author

## Acknowledgements

The authors would like to thank Prof. Maigeng Zhou and Dr. Yingying Jiang from Chinese Center for Disease Control and Prevention and Prof. Jiangmei Qin from China National Health Development Research Centre for their support to this project.

## Reference

1. Mökander J, Juneja P, Watson DS, Floridi L. The US Algorithmic Accountability Act of 2022 vs. The EU Artificial Intelligence Act: what can they learn from each other? Minds and Machines 2022: 1–8.

2. Möhlmann M, Zalmanson L, Henfridsson O, Gregory RW. Algorithmic Management of Work on Online Labor Platforms: When Matching Meets Control. MIS Quarterly 2021; 45(4): 1999–2022.

3. Cheng MM, Hackett RD. A critical review of algorithms in HRM: Definition, theory, and practice. Human Resource Management Review 2021; 31(1): 100698.

4. Weiskopf R, Hansen HK. Algorithmic governmentality and the space of ethics: Examples from ‘People Analytics’. Human Relations 2023; 76(3): 483–506.

5. Hassard J, Morris J. The extensification of managerial work in the digital age: Middle managers, spatio-temporal boundaries and control. Human Relations 2021; 0(0): 00187267211003123.

6. Mäntymäki M, Baiyere A, Islam AKMN. Digital platforms and the changing nature of physical work: Insights from ride-hailing. International Journal of Information Management 2019; 49: 452–60.

7. Parviainen P, Tihinen M, Kääriäinen J, Teppola S. Tackling the digitalization challenge: how to benefit from digitalization in practice. International journal of information systems and project management 2017; 5(1): 63–77.

8. Brennen JS, Kreiss D. Digitalization. The international encyclopedia of communication theory and philosophy 2016: 1–11.

9. Caza BB, Reid EM, Ashford SJ, Granger S. Working on my own: Measuring the challenges of gig work. Human Relations 2021; 75(11): 2122–59.

10. Denzin NK, Lincoln YS. The Sage handbook of qualitative research. Fifth ed. London: SAGE; 2017.

11. Wei H, Daniels S, Whitfield CA, et al. Agility and Sustainability: A Qualitative Evaluation of COVID-19 Non-pharmaceutical Interventions in the UK Logistics Sector. Front Public Health 2022; 10.

12. Möhlmann M, Zalmanson L. Hands on the wheel: Navigating algorithmic management and Uber drivers’. Autonomy’, in proceedings of the international conference on information systems (ICIS), Seoul South Korea; 2017; 2017. p. 10–3.

13. Veen A, Barratt T, Goods C. Platform-Capital’s ‘App-etite’ for Control: A Labour Process Analysis of Food-Delivery Work in Australia. *Work*, Employment and Society 2020; 34(3): 388–406.

14. Gandini A. Labour process theory and the gig economy. Human Relations 2019; 72(6): 1039–56.

15. Chen JY, Qiu JL. Digital utility: Datafication, regulation, labor, and DiDi’s platformization of urban transport in China. Chinese Journal of Communication 2019; 12(3): 274–89.

16. Zhang C, Cheung SP, Huang C. Job Demands and Resources, Mindfulness, and Burnout Among Delivery Drivers in China. Front Psychol 2022; 13.

17. Xie Y, Tian J, Jiao Y, Liu Y, Yu H, Shi L. The Impact of Work Stress on Job Satisfaction and Sleep Quality for Couriers in China: The Role of Psychological Capital. Front Psychol 2021; 12.

18. Wang Z, Neitzel RL, Zheng W, Wang D, Xue X, Jiang G. Road safety situation of electric bike riders: A cross-sectional study in courier and take-out food delivery population. Traffic Injury Prevention 2021; 22(7): 564–9.

19. Li J. Karoshi: An international work-related hazard? International Journal of Cardiology 2016; 206: 139–40.

20. Lin Y 林, Li Y 李. 心理资本、组织支持感在职业紧张与过度劳动关系中的作用—基于北京地区外卖骑手的调查 (The Role of Psychological Capital and Organizational Support in the Relationship between Occupational Stress and Overwork—Based on a Survey of Take-out Riders in Beijing). 中国流通经济(China Business And Market) 2021; 35(4): 116–26.

21. Lin Y 林, Li X 李, Li Y 李. 北京市快递员过劳现状及其影响因素—基于 1214 名快递员的调查 (Empirical Research on Employment and Overwork Situation of Couriers—Based on the Investigation of 1214 Couriers in Beijing). 中国流通经济(China Business And Market) 2018; 32(08): 79–88.

22. Zhang C, Sitar S, Huang C-C. Effects of Job Demands and Resources on Positive and Negative Affect of Delivery Drivers in China. International Journal of Environmental Research and Public Health 2022; 19(13): 8140.

23. Hong Y, Zhang Y, Xue P, et al. The Influence of Long Working Hours, Occupational Stress, and Well-Being on Depression Among Couriers in Zhejiang, China. Front Psychol 2022; 13.

24. Lee J, Di Ruggiero E. How does informal employment affect health and health equity? Emerging gaps in research from a scoping review and modified e-Delphi survey. International Journal for Equity in Health 2022; 21(1): 87.

25. Guan M. Should the poor have no medicines to cure? A study on the association between social class and social security among the rural migrant workers in urban China. International Journal for Equity in Health 2017; 16(1): 193.

26. Cram WA, Wiener M, Tarafdar M, Benlian A. Examining the Impact of Algorithmic Control on Uber Drivers’ Technostress. Journal of Management Information Systems 2022; 39(2): 426–53.

27. Sun G. Quantitative Analysis of Online Labor Platforms’ Algorithmic Management Influence on Psychological Health of Workers. International Journal of Environmental Research and Public Health 2023; 20(5): 4519.

28. Rani U, Pesole A, Gonzalez Vazquez I. Algorithmic Management practices in regular workplaces: case studies in logistics and healthcare. Luxembourg: Publications Office of the European Union, 2024.

29. Meijerink J, Bondarouk T. The duality of algorithmic management: Toward a research agenda on HRM algorithms, autonomy and value creation. Human Resource Management Review 2021: 100876.

30. Schuetz S, Venkatesh V. Research Perspectives: The Rise of Human Machines: How Cognitive Computing Systems Challenge Assumptions of User-System Interaction. Journal of the Association for Information Systems 2020; 21(2).

31. Ivanova M, Bronowicka J, Kocher E, Degner A. The App as a Boss? Control and Autonomy in Application-Based Management. 2018.

32. Fairclough N. Peripheral Vision: Discourse Analysis in Organization Studies: The Case for Critical Realism. Organization Studies 2005; 26(6): 915–39.

33. Fleiss JL, Levin B, Paik MC. Statistical methods for rates and proportions: john wiley & sons; 2013.

34. Tan ZM, Aggarwal N, Cowls J, Morley J, Taddeo M, Floridi L. The ethical debate about the gig economy: A review and critical analysis. Technology in Society 2021; 65: 101594.

35. Karanović J, Berends H, Engel Y. Regulated Dependence: Platform Workers’ Responses to New Forms of Organizing. Journal of Management Studies 2021; 58(4): 1070–106.

36. Gal U, Jensen TB, Stein M-K. Breaking the vicious cycle of algorithmic management: A virtue ethics approach to people analytics. Information and Organization 2020; 30(2): 100301.

37. Zhang A, Boltz A, Wang CW, Lee MK. Algorithmic Management Reimagined For Workers and By Workers: Centering Worker Well-Being in Gig Work. Proceedings of the 2022 CHI Conference on Human Factors in Computing Systems. New Orleans, LA, USA: Association for Computing Machinery; 2022. p. Article 14.

38. Heiland H. Neither timeless, nor placeless: Control of food delivery gig work via place-based working time regimes. Human Relations 2022; 75(9): 1824–48.

39. Le Breton C, Galière S. The role of organizational settings in social learning: An ethnographic focus on food-delivery platform work. Human Relations 2023; 76(7): 990–1016.

40. Li AK. Beyond algorithmic control: flexibility, intermediaries, and paradox in the on-demand economy. Information, communication & society 2022; 25(14): 2012–27.

41. Zhang L, Yang J, Zhang Y, Xu G. Gig worker’s perceived algorithmic management, stress appraisal, and destructive deviant behavior. PLOS ONE 2023; 18(11): e0294074.

42. Prabowo R, Sucahyo YG, Gandhi A, Ruldeviyani Y. Does Gamification Motivate Gig Workers? A Critical Issue in Ride-Sharing Industries. 2019 International Conference on Advanced Computer Science and information Systems (ICACSIS); 2019 12-13 Oct. 2019; 2019. p. 343–8.

